# Warming temperatures could expose more than 1.3 billion new people to Zika virus risk by 2050

**DOI:** 10.1101/2020.06.29.20142422

**Authors:** Sadie J. Ryan, Colin J. Carlson, Blanka Tesla, Matthew H. Bonds, Calistus N. Ngonghala, Erin A. Mordecai, Leah R. Johnson, Courtney C. Murdock

## Abstract

In the aftermath of the 2015 pandemic of Zika virus, concerns over links between climate change and emerging arboviruses have become more pressing. Given the potential that much of the world might remain at risk from the virus, we use a model of thermal bounds on Zika virus (ZIKV) transmission to project climate change impacts on transmission suitability risk by mid-century (a generation into the future). In the worst-case scenario, over 1.3 billion new people could face suitable transmission temperatures for ZIKV by 2050. Given these suitability risk projections, we suggest an increased priority on research establishing the immune history of vulnerable populations, modeling when and where the next ZIKV outbreak might occur, evaluating the efficacy of conventional and novel intervention measures, and increasing surveillance efforts to prevent further expansion of ZIKV.

**Author Summary:** First discovered in Uganda in the 1950s, Zika virus (ZIKV) is a new threat to global health security. The virus is spread primarily by female *Aedes* mosquitoes, with occasional sexual transmission in humans, and can cause Zika congenital syndrome (which includes fetal abnormalities like microcephaly) when women are infected during pregnancy. Our study is the first to quantify how many people may be exposed to temperatures suitable for ZIKV transmission in a changing climate. In the worst-case scenario, by 2050, climate change could expose more than 1.3 billion people worldwide to temperatures suitable for transmission - for the first time. The next generation will face substantially increased ZIKV transmission temperature suitability in North America and Europe, where naïve populations might be particularly vulnerable. Mitigating climate change even to moderate emissions scenarios could significantly reduce global expansion of climates suitable for ZIKV transmission, potentially protecting around 200 million people.

## Introduction

In the past two decades, several emerging arboviruses have undergone continental or global range expansions. Often, these viruses only become major research targets after sizable outbreaks in the Western hemisphere, like the 1999-2002 West Nile virus outbreak in the United States and Canada [1], the 2013-2015 chikungunya outbreak in the Americas, and, most recently, the 2015-2017 epidemic of Zika virus (ZIKV) in the Americas. Although these outbreaks often take public health communities by surprise [1, 2], their international spread occurred gradually with limited global concern and little action before a major outbreak occurred [4]. In the aftermath of the Zika pandemic, developing tools that can successfully anticipate the climate conditions that promote these sorts of explosive arbovirus outbreaks is a critical research need [4, 5].

ZIKV continues to pose a looming threat. Since 2013, the virus spread to at least 49 countries and territories [7], resulting in an estimated 150,000 to 500,000 cases with at least 3,000 cases of microcephaly in Brazil alone [3]. While other emerging and re-emerging viruses have much higher case-fatality rates [8], such as yellow fever virus or Eastern equine encephalitis virus, the social and human costs of microcephaly are profound [9]. In the aftermath of the epidemic in the Americas [6, 9], concern remains about potential future outbreaks of the virus and its ongoing relevance as a public health threat. In Latin America and the Caribbean, high seroprevalence rates would suggest that another major outbreak is unlikely in the short term [11, 12]. In contrast, ZIKV remains a potential threat at its range margins in the Americas (especially in the southern United States) [13, 14], Africa, and Asia [15–17]. Although the virus has spread through Africa and Asia for several decades without an outbreak on the scale of the one in the Americas, the seroprevalence of Zika in its native range is poorly characterized. Further, the evolution of novel ZIKV strains may make the threat newly relevant in Africa and Asia [18]: local transmission of ZIKV in Angola in 2017 underscores the potential for this problem to occur again [19].

The relationship between climate change and ZIKV adds an additional layer of complexity. The contribution of climate change to the severity of the 2015 outbreak is difficult to ascertain definitively, and some have suggested links between Zika transmission and El Niño [20]. Previous models have suggested that *Aedes*-borne virus transmission should expand significantly in a changing climate, especially those transmitted by *Aedes aegypti* [21]. Limited modeling work done during the 2016 outbreak suggested that Zika transmission might be constrained to slightly warmer, less seasonally variable parts of the world than dengue [22]; and recent work by Tesla *et al*. combining experimental and modeling approaches has suggested that the minimum temperature for ZIKV transmission by *Aedes aegypti* is roughly 5°C higher than that of dengue virus [23]. Thus, while the current range of Zika transmission is confined to the tropics, climate change could increase the number of people exposed for the first time to temperatures suitable for Zika transmission.

Here, we provide the first systematic assessment of where future temperatures are expected to become suitable for transmission, and could most substantially increase the distribution of ZIKV and its risk to human populations. To achieve this, we follow a similar approach described in our previous studies that have used a temperature-dependent transmission model to assess environmental suitability for dengue transmission [24]. In this study, we project the Zika-specific model [23] onto current temperatures and evaluate the population at risk based on human population density data from 2015, during the Zika pandemic. We then project the temperature-dependent Zika model forward with climate change to the year 2050 (approximately one human generation into the future) and evaluate where human populations might be expected to face their first exposure to temperatures suitable for ZIKV transmission.

## Methods

### Mechanistic Model of Temperature Suitability for ZIKV Transmission

We used a recently published experimentally-derived mechanistic model of ZIKV transmission by *Ae. aegypti* to map temperature-driven transmission risk [23]. In brief, the approach is to use a Bayesian framework to fit thermal responses for mosquito and virus traits that drive transmission that were empirically estimated in laboratory experiments, and then combine them to obtain the posterior distribution of *R*_*0*_ as a function of temperature. The full methods are described in detail in Johnson *et al*. [25] and several of the particular traits and fits for *Ae. aegypti* are originally presented in Mordecai *et al*. [26]. The more recent Tesla *et al*. description of thermal responses included data and fitted thermal performance curves for daily adult mosquito mortality as well as two ZIKV specific traits: vector competence and the extrinsic incubation rate [23]. The posterior samples for *R*_*0*_ as a function of temperature (rescaled to range from zero to one, given that the absolute magnitude of *R*_*0*_ in any given setting varies) were generated, and the probability that *R*_*0*_ > 0 at each temperature was obtained, a cutoff inclusive of any transmission risk (not just sustained outbreaks, where *R*_*0*_ > 1). We used the thermal boundaries for which ZIKV *R*_*0*_ > 0 with a posterior probability > 0.975 to define the limits on suitability for transmission for monthly temperatures, then calculated climate model data layers as described below. This high probability allows us to define a temperature range for potential transmission that is conservative. The final interval of “suitable transmission temperatures” was given as 23.9-34.0°C.

### Climate & Population Data

To examine the impact of climate change on transmission risk, we follow the approach of Ryan *et al*. [24]. Briefly, we obtained 5 minute resolution current mean monthly temperature data from the WorldClim dataset (www.worldclim.org) [27]. We then selected four general circulation models (GCMs) and two representative concentration pathways (RCPs 4.5, 8.5) to account for different global responses to mitigate climate change. The GCMs are the Beijing Climate Center Climate System Model (BCC-CSM1.1); the Hadley GCM (HadGEM2-AO and HadGEM2-ES); and the National Center for Atmospheric Research’s Community Climate System Model (CCSM4). Future scenario climate model output data were acquired from the research program on Climate Change, Agriculture, and Food Security (CCAFS) web portal (http://ccafs-climate.org/data_spatial_downscaling/), part of the Consultative Group for International Agricultural Research (CGIAR). We used model outputs created using the delta downscaling method, from the IPCC AR5. For visualizations, we used the HadGEM2-ES model, the most commonly used GCM. The mechanistic transmission model was then projected onto the climate data using the ‘raster’ package in R 3.1.1 (‘raster’ [28]).

To quantify the population at risk (PAR), we diverged from the previous approach, and aimed to not just capture projected population growth, but to incorporate influences of predicted economic and social changes during the next half century. Therefore, we updated the methods in Ryan *et al*. [21] to incorporate population projection products that contain the Shared Socioeconomic Pathways (SSPs) [29]. The SSPs are five alternative population trajectory outcomes based on development, economic, education, and urbanization trends, specifically tailored to responses to climate change and/or mitigation strategies. The different SSPs (1-5) describe trajectories in which components such as fertility and urban growth are impacted by differences in regional and national equality, conflict, efforts for sustainability, or driven by fossil fuels. Different combinations of SSPs and RCPs can be paired based on plausibility, but this introduces factorial combinations and makes coherent projections about future disease risk more complex, so we reserve exploration of this axis of demographic complexity for future studies. For this study, we selected the SSP2 scenario—a middle of the road scenario—to reflect expected growth and population geography shifts due to processes such as migration and urbanization. We acknowledge that SSP2 and RCP 8.5 are an unlikely combined future scenario [30,31], but present this as the extreme of our projected continuum. The population product we use here is projected from a baseline population, the Gridded Population of the World (GPW) [32], including the Global Rural-Urban Mapping Project (GRUMP) [33]. We therefore chose a baseline population for 2015 from GPW products, to reflect conditions during the recent Zika outbreak, and selected the SSP2 2050 population projection product [30], available from (http://sedac.ciesin.columbia.edu/data/set/popdynamics-pop-projection-ssp-2010-2100/data-download). We aggregated all geographic layers in our analyses to a 0.25° grid cell to be consistent.

### Current and Future Transmission Risk

To examine the impact of climate change on transmission risk, we follow the approach of Ryan *et al*. [21]. This previous work used existing *Aedes* transmission models (which are mostly appropriate for dengue) [26] to project climate change impacts, by mapping areas where mean temperatures fall within the 97.5% posterior probability, or 95% credibility interval for suitability predicted by the Bayesian model. These maps can be projected onto different climate futures (different general circulation models and representative concentration pathways; GCMs and RCPs, respectively), and populations at risk can be compared between current and future maps. Following this protocol, we overlay suitability maps and population grids for 2015 and 2050 (with different climate pathways for the latter) and calculate global population at risk. For each analysis, we also stratify these estimates with a regional breakdown using the definitions of the Global Burden of Disease (GBD) study regions to align with policy and planning goals [34].

## Results

At present, most of the predicted transmission risk for ZIKV occurs in the tropics (**Figure 1**). Using the Tesla *et al*. [23] thermal boundary projection map, we find a “population at risk” (PAR) for 2015 of ∼5 billion (here referring to any population inside pixels evaluated as thermally suitable for at least one month of the year – **Table 1**). It is important to note that this thermal boundary does not distinguish whether or not *Aedes aegypti* or ZIKV are currently present in a region. Outside Latin America and the Caribbean (LAC), we find a population at risk of 4.69 billion (mostly in South and East Asia). In contrast to the large proportion of the global population experiencing any temperature suitability for ZIKV, we find a total population-at-risk of 858 million (the vast majority, 767 million, outside LAC) in areas with year-round temperature suitability for transmission (pixels evaluated as thermally suitable for 12 months – **Table 2**), highlighting the locations with the most suitable climates where ZIKV importation could lead to sustained outbreaks.

**Figure 1.**
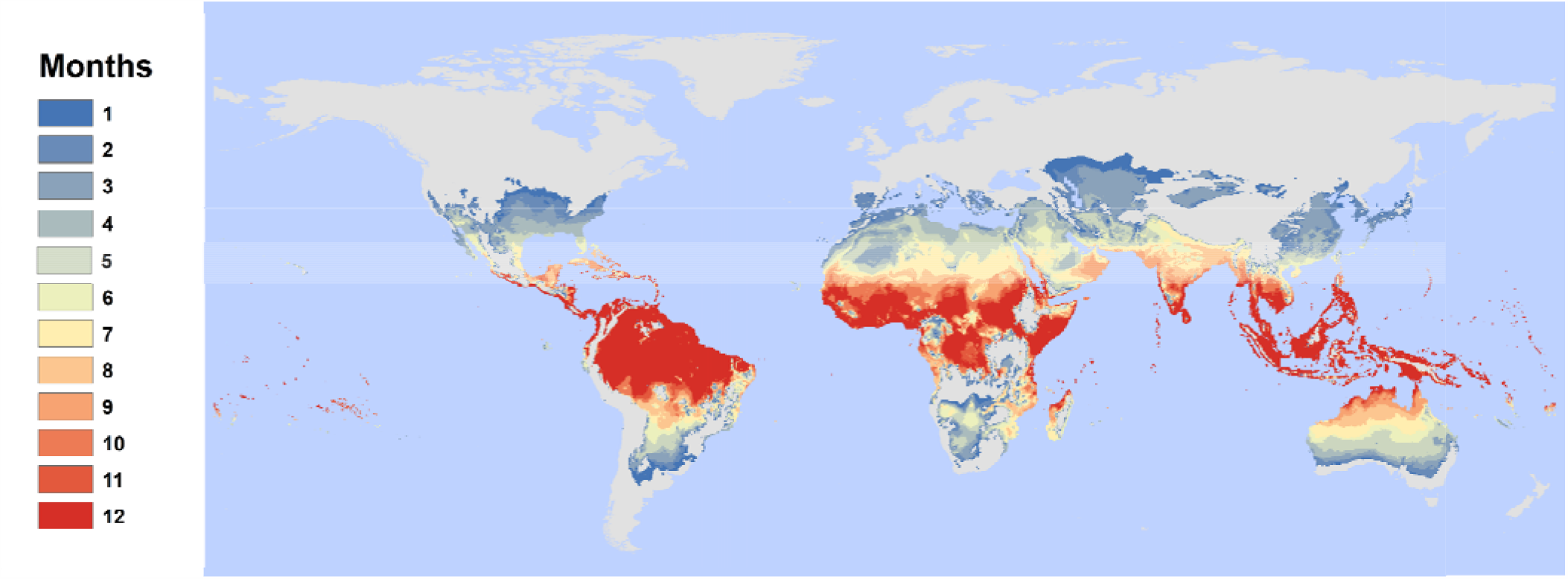
Current distribution of temperature suitability for Zika transmission, by month. Results show the number of suitable months per year based on a 97.5% posterior probability for *R*_*0*_(*T*) > 0 based on the Tesla *et al*. (2018) model of Zika transmission, as a function of mean monthly temperature in each pixel.

**Table 1.** Current and projected net changes in population at risk for any transmission (one or more months). All values are given in millions; future projections are averaged across general circulation models (GCMs), broken down by year (2050) and representative concentration pathway (RCP: 4.5, 8.5), and are given as net change from a baseline of 2015 population at risk. Totals are given globally, or across all regions except for Latin America and the Caribbean (LAC).

| Region | 2015 | 2050 |  |
| --- | --- | --- | --- |
|  |  | RCP 4.5 | RCP 8.5 |
| Asia (Central) | 56.5 | 20.3 | 24.8 |
| Asia (East) | 1,113.5 | 50.3 | 83.8 |
| Asia (High Income Pacific) | 138.1 | -5.8 | -4.2 |
| Asia (South) | 1,621.9 | 639.2 | 644.7 |
| Asia (Southeast) | 520.7 | 133.7 | 142.5 |
| Australasia | 5.3 | 11.4 | 14.5 |
| Caribbean | 33.8 | 2.2 | 2.7 |
| Europe (Central) | 0.6 | 52.2 | 67.7 |
| Europe (Eastern) | 5.0 | 69.5 | 91.0 |
| Europe (Western) | 42.9 | 106.9 | 131.2 |
| Latin America (Andean) | 17.3 | 15.1 | 16.0 |
| Latin America (Central) | 110.8 | 66.1 | 76.0 |
| Latin America (Southern) | 15.1 | 30.8 | 31.9 |
| Latin America (Tropical) | 119.7 | 56.8 | 67.0 |
| North Africa & Middle East | 359.6 | 262.9 | 275.3 |
| North America (High Income) | 163.9 | 195.9 | 213.8 |
| Oceania | 4.2 | 4.0 | 4.2 |
| Sub-Saharan Africa (Central) | 86.1 | 100.2 | 106.3 |
| Sub-Saharan Africa (East) | 179.8 | 315.3 | 344.9 |
| Sub-Saharan Africa (Southern) | 14.3 | 29.4 | 37.3 |
| Sub-Saharan Africa (West) | 373.8 | 340.4 | 341.0 |
| **Total** | **4,928.7** | **2,496.8** | **2,712.5** |
| **Total outside LAC** | **4,686.0** | **2,325.8** | **2,518.9** |

**Table 2.** Current and projected net changes for population at risk from year-round transmission risk (12 months). All values are given in millions; future projections are averaged across GCMs, broken down by year (2050) and RCP (4.5, 8.5), and are given as net change from current population at risk. Totals are given globally, or across all regions except for Latin America and the Caribbean (LAC).

| Region | 2015 | 2050 |  |
| --- | --- | --- | --- |
|  | Baseline | RCP 4.5 | RCP 8.5 |
| Asia (Central) | 0 | 0 | 0 |
| Asia (East) | 0 | 0 | 0.005 |
| Asia (High Income Pacific) | 2.7 | 1.5 | 1.4 |
| Asia (South) | 104.5 | 111.4 | 107.6 |
| Asia (Southeast) | 356.7 | 164.0 | 179.0 |
| Australasia | 0.1 | 0.1 | 0.1 |
| Caribbean | 7.5 | 15.2 | 19.1 |
| Europe (Central) | 0 | 0 | 0 |
| Europe (Eastern) | 0 | 0 | 0 |
| Europe (Western) | 0 | 0 | 0 |
| Latin America (Andean) | 5.2 | 8.8 | 10.2 |
| Latin America (Central) | 43.7 | 44.3 | 55.0 |
| Latin America (Southern) | 0 | 0 | 0 |
| Latin America (Tropical) | 35.0 | 16.5 | 23.2 |
| North Africa & Middle East | 5.0 | -1.3 | -0.01 |
| North America (High Income) | 0 | 0.001 | 0.01 |
| Oceania | 2.5 | 3.3 | 3.8 |
| Sub-Saharan Africa (Central) | 28.0 | 92.0 | 106.6 |
| Sub-Saharan Africa (East) | 39.7 | 106.8 | 148.3 |
| Sub-Saharan Africa (Southern) | 0 | 0 | 0 |
| Sub-Saharan Africa (West) | 227.4 | 281.8 | 261.5 |
| **Total** | **857.9** | **844.3** | **915.9** |
| **Total outside LAC** | **766.5** | **759.5** | **808.3** |

We predict that unmitigated climate change could shift as many as 1.33 billion new people (1.17 billion outside LAC) into areas with future temperatures suitable for ZIKV transmission under the worst-case scenario (RCP 8.5; **Table 3**). Five regions with populations of 100 million or more people are projected to experience climate suitability for Zika transmission: East Africa, High-income North America, East Asia, Western Europe, and North Africa and the Middle East (with regions designated by the Global Burden of Disease study). A total of 737 million people worldwide (635.8 million outside LAC) could face their first exposure to year-round climate suitability for Zika transmission, mostly in South and East Asia and sub-Saharan Africa.

**Table 3.**
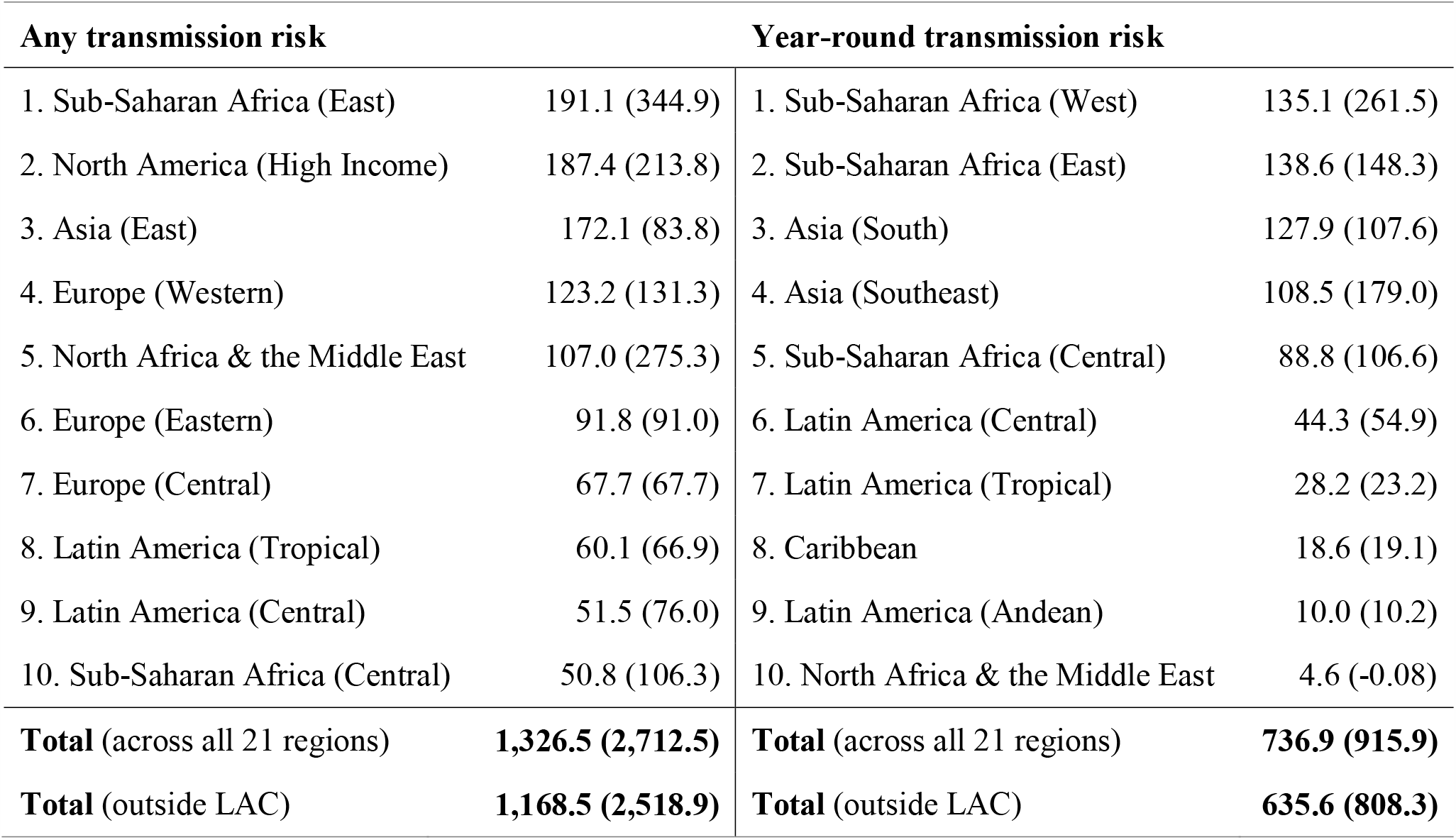
Top 10 regional increases. Regions, as defined by the Global Burden of Disease study (Fig. S1) are ranked based on millions of people exposed for the first time to any (1 or more months) transmission risk, or to year round (12 months) transmission risk; parentheticals give the net change (first exposures minus populations escaping transmission risk). All values are given for the worst-case scenario (RCP 8.5). Totals are given globally, or across all regions except for Latin America and the Caribbean (LAC).

Net changes in risk are dramatic, largely because there are very few areas where climate warming will drive future temperatures to become unsuitable (too hot) for at least one month of the year, but many areas where the climate will become newly suitable (**Figure 2**). For any transmission risk suitability (**Table 1**), we find a net increase of 2.71 billion (2.52 billion outside LAC) people at risk in the worst-case scenario (RCP 8.5). Even in the more moderate scenario for climate change mitigation (RCP 4.5), we project a net increase of 2.50 billion people at risk (2.33 billion outside LAC; **Table 1**). For people living in areas that experience year-round risk (**Table 2**), we project for the moderate- and worst-case scenarios a minimum increase of 844.3 million (759.5 million outside LAC) and 915.9 million (808.3 million outside LAC), respectively. Therefore, the majority of net changes in people at risk for ZIKV-suitable climates occurs even under the partial mitigation (RCP 4.5) scenario.

**Figure 2.**
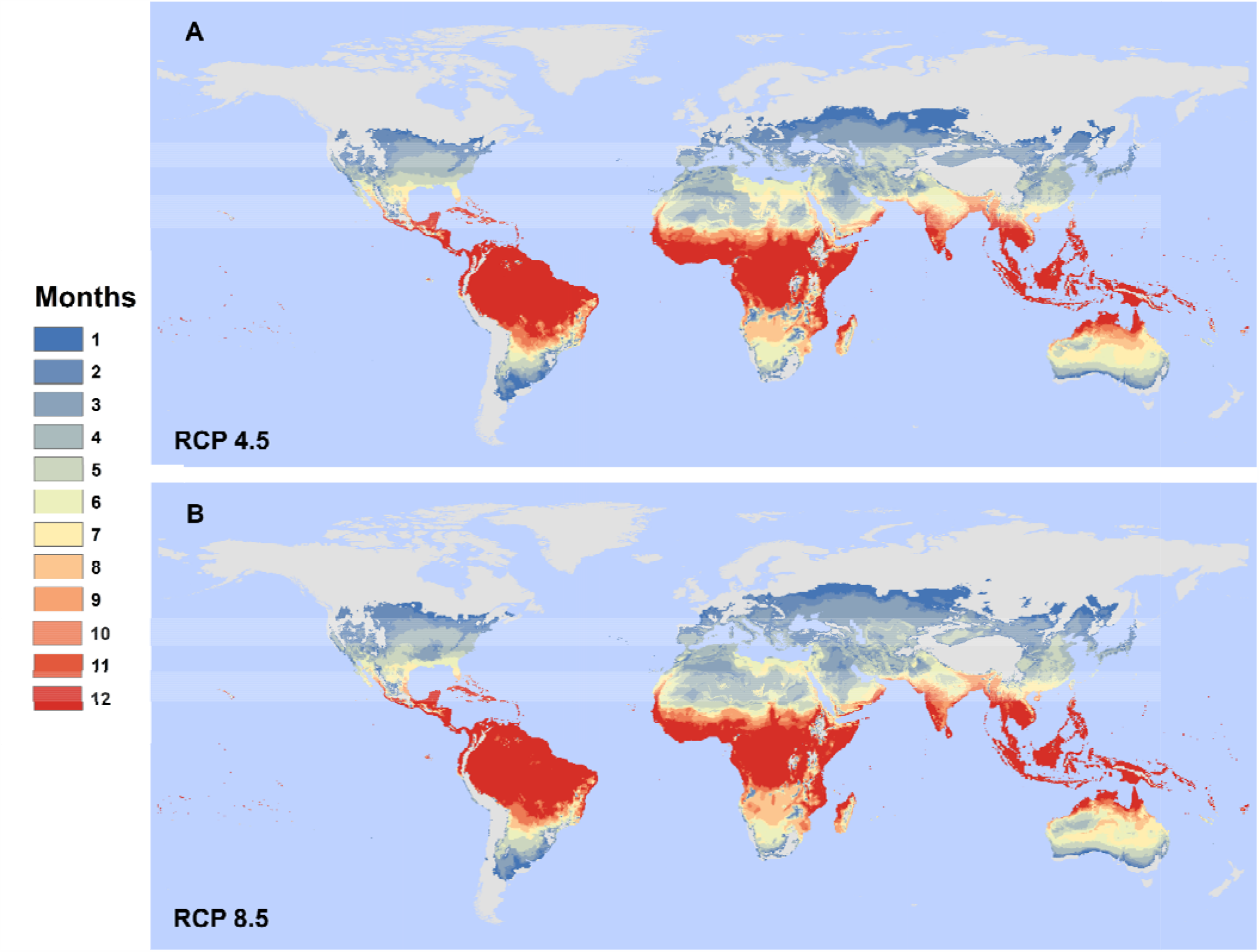
A moderate-case and worst-case scenario for 2050. The figure shows our model projecting Zika transmission risk for (A) RCP 4.5 and (B) RCP 8.5 (HadGEM2-ES). Results show the number of suitable months per year based on a 97.5% posterior probability for *R*_*0*_(*T*) > 0 based on the Tesla *et al*. (2018) model of Zika transmission, as a function of mean monthly temperature in each pixel.

In the moderate-case scenario (RCP 4.5), the region experiencing the largest increase in first exposures to any (one or more months) transmission suitability is high income North America (169.5 million), while under the worst-case scenario (RCP 8.5), the top region is Eastern sub-Saharan Africa (191.1 million); these regional increases are shown in **Figure 3**.

**Figure 3.**
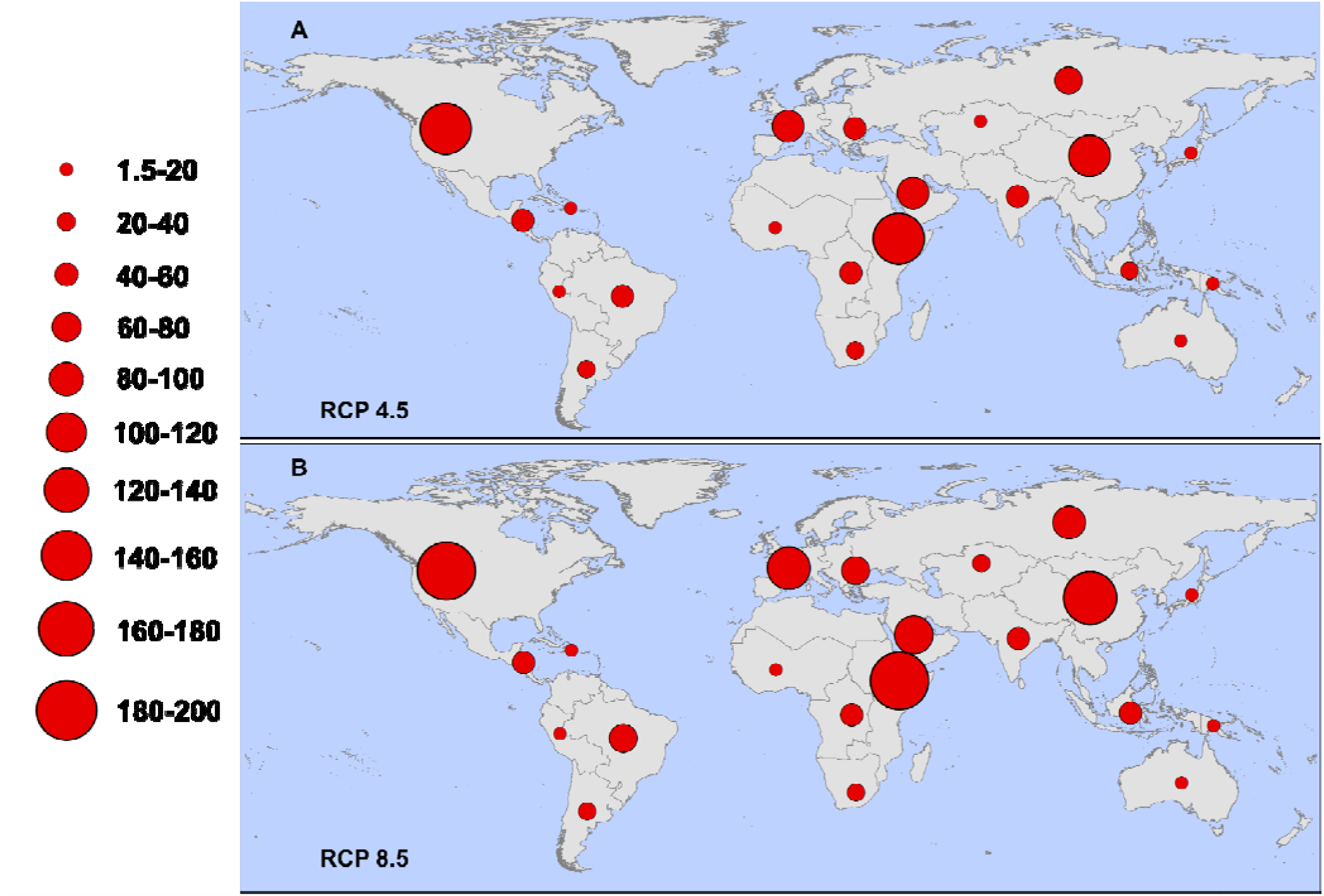
Regional increases in populations at risk for any transmission (one or more months). Regions are defined according to the Global Burden of Disease (GBD) regions (detailed in Figure S1), and proportional red circles illustrate the regional populations (in millions) at risk under (A) RCP 4.5 and (B) RCP 8.5.

## Discussion

We present an upper bound on potential future expansion of ZIKV transmission risk based on thermal suitability in a changing climate. When compared to mapped projections for dengue transmission suitability in *Aedes aegypti* [24], we see a more constrained range, as the lower temperature limit for ZIKV transmission is higher, precluding cooler regions. This difference in predicted spatial suitability corroborates findings from distribution modeling approaches to human case data for dengue and ZIKV [22], underscoring the importance of understanding transmission biology of specific vector-pathogen pairings. Our mechanistic, trait-based modeling approach has successfully explained the distribution of several vector-borne illnesses, including dengue [26], malaria [35], and Ross River fever [36].

Our results indicate that warming temperatures will increase thermal suitability for ZIKV transmission in a significant portion of the world, with over 1.3 billion people likely to be exposed for the first time to temperature conditions suitable for Zika transmission by the mid-century in the worst-case scenario. In combination with population change in at-risk areas, this produces an increase in the population at risk on the order of 2.7 billion people. Whether this risk leads to actual re-emergence events depends on several limiting factors, most of all the presence or absence of competent mosquito vectors. The Zika transmission model presented here already designates some areas as suitable that are outside the range of *Ae. aegypti*, the main vector, or *Ae. albopictus*, another competent vector of concern. However, both *Ae. aegypti* and *Ae. albopictus* are projected to expand their range to higher latitudes and elevations, with some indication this has already happened [37,38]. Other studies have also highlighted the potential for ZIKV to be transmitted by other mosquitoes that are widespread in the rest of the predicted range [39–41]; each new mosquito vector species and its relative contribution to potential transmission would introduce subtle differences in the realized climate suitability and effects of climate change on ZIKV transmission [21,26]. Even where conditions are suitable and mosquito vectors are present, repeated importation may not lead to establishment [42,43] due to a combination of stochasticity and confounding and interconnected socioenvironmental risk factors (e.g., housing construction, water storage, mosquito control, and surveillance efforts) [44,45].

As the risk areas for ZIKV expand, prioritizing regions for intervention becomes more difficult. Several major research advances can improve predictions. Testing the competence of *Aedes* mosquitoes and others to transmit the virus, and identifying regional differences in competence, are key steps [40,41,46–50]. The impacts of the immune history and genetic risk factors of human populations are an additional critical component of the system [51]. Seroprevalence studies indicate that another catastrophic Zika epidemic is unlikely in Latin America and the Caribbean in the near term [11,12], but much less is known about ZIKV susceptibility in African, Asian, and Pacific Islander populations. Similarly, microcephaly rates in response to ZIKV infection varied even within the Americas, with the highest rates in Brazil [52] and lower rates in the Caribbean [53] suggesting significant variation exists across human populations in the severity of symptoms associated with ZIKV infection. Given the potential for explosive outbreaks in naïve populations (as happened in the Americas), this is a top priority for predicting the potential for future outbreaks. The relevance of ZIKV in the coming decades will be determined by the overall risk of microcephaly, Zika congenital syndrome more broadly, and Guillain-Barre syndrome—the most severe manifestations of ZIKV infection—which remain poorly understood.

Other features of the abiotic environment, like precipitation, relative humidity, or solar radiation, also constrain the distribution of vectors and their pathogens, as do biotic interactions and human-modified features of the landscape. Some of these factors impact transmission in ways that can be surprising. For example, typically it is assumed that increased precipitation should increase arboviral transmission, due to increasing potential larval habitat. However, during the 2015-2017 ZIKV outbreak, there was an inverse relationship with precipitation: drought led to greater transmission because increases in household water storage were associated with increased Zika cases [44,54] (a pattern also observed in previous arbovirus outbreaks [55]). Importantly, as mosquitoes are ectothermic, and the effects of precipitation on transmission are not straightforward, using thermal suitability allows us to set range boundaries on future risk, both geographically and seasonally. Moreover, in addition to average temperature suitability, temperature variation can play an important and nonlinear role in transmission, at the level of the vector [56], potentially increasing environmental suitability for transmission at low mean temperatures. In addition to this, warming in areas with higher mean temperatures is predicted to reduce suitability, as conditions pass beyond optimal transmission suitability temperatures. This can lead to reductions in predicted risk, as seen in the high income Pacific Asia region (Table 1); this echoes findings for malaria suitability in Africa, seen in Ryan et al [57,58], wherein Western Africa becomes too hot for malaria suitability, and risk appears to decline rapidly under climate change scenarios. Given the high upper thermal bounds of transmission suitability of ZIKV, other direct or indirect impacts of heat on human health are likely to arise, meaning these predicted declines at high temperatures are not necessarily an optimistic picture. Our temperature-based approach isolates one of the strongest environmental filters, and avoids confounding issues of the relationship between precipitation and arboviruses that occurs in the urban environment, where human behavior and water practices may drive dynamics; put simply, for urban arboviral transmission, where people are, so is water. Hopefully, the future risk we project in this study is likely to be substantially constrained by limiting protective factors from vector-borne disease infection, including socioeconomic, immunological, intervention, and environmental factors, including the built environment itself.

The ZIKV outbreak originating in Brazil in 2015 quickly became a historically significant global health emergency, highlighting the growing threat of emerging diseases in a changing world [55]. Even in the moderate-case mitigation scenario (RCP 4.5) considered here, climate change will substantially increase climate suitability for Zika outbreaks in tropical and temperate zones around the world. With the economic and social costs of the 2015-2017 pandemic still accumulating, our results suggest another case in which climate change mitigation is unequivocally necessary for the sake of global health security.

## Data Availability

Mapped results will be available through Harvard Dataverse at publication acceptance through URL XXX-XXX

## Author contributions

SJR, CJC, EAM, CCM, LRJ, and BT designed the modeling and analysis frameworks. CJC and SJR performed the analyses and wrote the first draft. All authors contributed to the writing and design of the paper and approved the final submission.

## Acknowledgements

Thanks to Fausto Bustos for critical comments on Zika immunology. We would also like to thank Dr. Melinda Brindley and Leah Demakovsky in the design and execution of the study that generated the Zika virus data that informed our temperature-dependent *R*_*0*_ model.

## Supplementary

**S1 Figure.**
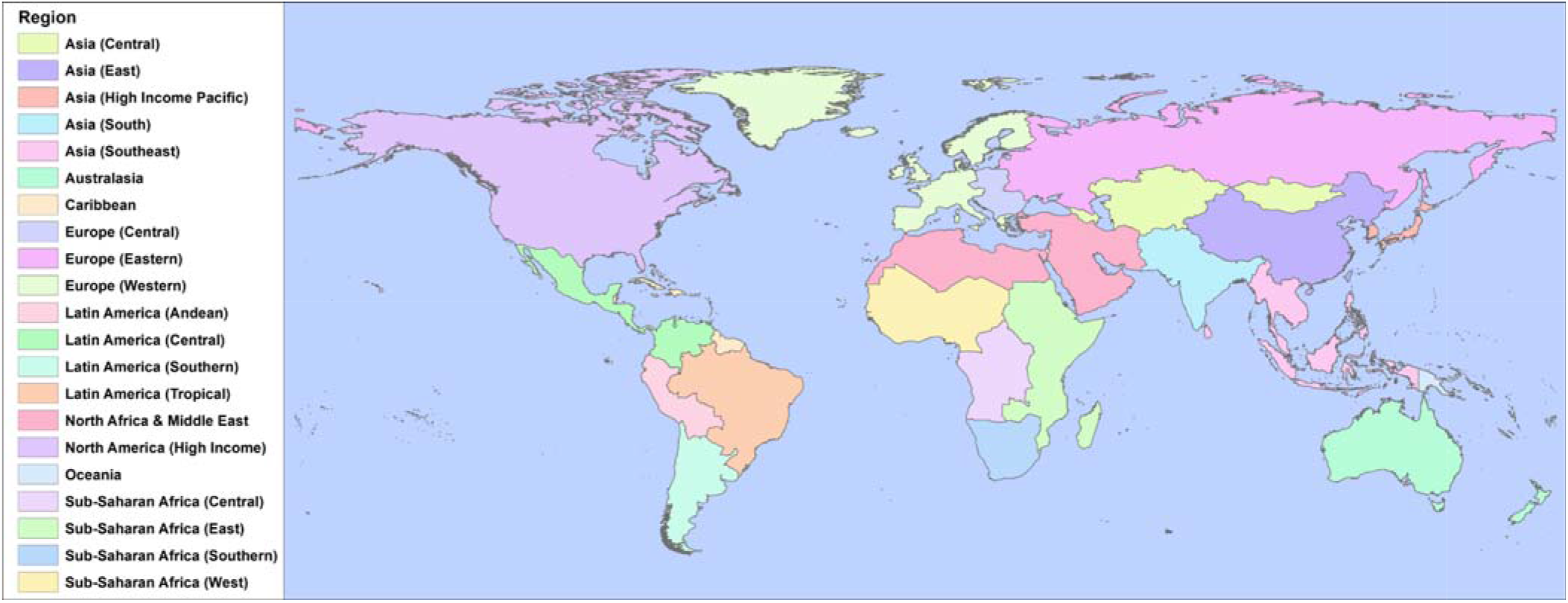
Global health regions. We adopt the same system as the Global Burden of Disease Study in our regional breakdown.

